# Major UK non-commercial sponsors’ efforts to reduce research waste: a mixed methods study

**DOI:** 10.1101/2023.05.03.23289434

**Authors:** Till Bruckner, Aminul Schuster, Belén Chavarría, Carolina Cruz, Karely Lizárraga, Ronak Borana, Tungamirai Ishe Bvute, Daniel Sánchez

## Abstract

This study provides a snapshot of the scale of legacy research waste in the UK. It assesses the current publication status of 145 clinical trials sponsored by ten major UK non-commercial sponsors that were completed or terminated in 2017. Following outreach to sponsors and short-term follow-up, 116/145 trials (80%) had fully reported results, and 11/145 trials (8%) had reported results in the grey literature. Results for 18/145 trials (12%) that enrolled 637 people remained completely unpublished as of early March 2023. Sponsors indicated that they plan to make public the results of 14/18 unreported trials. Our study had an impact on accelerating the reporting of some results, and seems likely to lead to future reductions in research waste. We propose three changes to UK Health Research Authority policies that could improve clinical trial reporting.

## BACKGROUND

Worldwide, a significant proportion of clinical trials end up as costly research waste because their results are never made public. The resulting gaps in the medical evidence base harm patients and undermine public health.^1^

In the wake of a 2018 UK parliamentary enquiry and sustained engagement by advocacy groups and UK public bodies, non-commercial clinical trial sponsors in the UK substantially improved outcome reporting for drug trials (Clinical Trials of Investigative Medicinal Products, CTIMPs) by uploading the summary results of many CTIMPs onto the European Union Clinical Trials Register (EUCTR), including for older legacy trials.^2^ However, previous research indicates that many institutions’ efforts to improve trial reporting did not extend to other types of trials listed on other trial registries.^3^ The UK’s national #MakeItPublic strategy now aims to ensure that going forward, all clinical trials involving UK patients will make their results public, but the strategy’s scope does not extend retrospectively to older legacy trials.^4^

Previous studies have found that in the absence of any external intervention, the results of very few trials that remain unpublished after 5 years will ever be made public.^5^ However, a recent TranspariMED project in Germany indicated that outreach to institutions that sponsored trials that have remained unreported in the past can spur non-commercial sponsors to tackle legacy research waste.^6^

This study assesses the current publication status of 145 clinical trials sponsored by ten major UK non-commercial sponsors that were completed or terminated in 2017, before the launch of the parliamentary enquiry, in order to provide a snapshot of the scale of legacy research waste in the UK.

The Declaration of Helsinki, which stipulates that all clinical trial results must be made public, albeit without specifying the reporting format or timeframe, is applicable to all trials in the study cohort. ^7^ However, there is no legal requirement for UK sponsors to make trial results public.

## METHODOLOGY

### Cohort selection

The lead researcher (TB) identified the ten most prolific non-commercial sponsors of clinical trials in the UK by accessing the EU Trials Tracker on 27 October 2022, employing the number of drug trials (CTIMPs) run by each sponsor as a proxy indictor of overall trial volume.^8^

**Table 1:** Selection of UK sponsors according to total number of CTIMPs sponsored

| Sponsor | CTIMPs | With results* |
| --- | --- | --- |
| University College London | 163 | 97% |
| University of Oxford | 146 | 99% |
| Imperial College London | 145 | 97% |
| University of Birmingham | 118 | 100% |
| King's College London | 107 | 99% |
| Guy's and St Thomas' NHS Foundation Trust | 82 | 94% |
| University of Dundee | 74 | 100% |
| University of Leeds | 72 | 98% |
| NHS Greater Glasgow and Clyde | 71 | 100% |
| Newcastle upon Tyne Hospitals NHS [Foundation] Trust | 69 | 93% |

*\* Due trial results posted on EUCTR, as per EU Trials Tracker data*

The lead researcher then used the advanced search functions of the two other trial registries commonly used by UK sponsors, ClinicalTrials.gov and ISRCTN, on 28 October 2022 to identify all clinical trials run by these ten sponsors that were completed or terminated in 2017. Inclusion criteria:

- The (lead) sponsor was one of the ten UK non-commercial sponsors listed above
- Interventional clinical trial completed or terminated 01 January 2017 and 31 December 2017

The lead researcher applied these search criteria to both registries and extracted the trial ID numbers, patient enrolment numbers, and registry reporting status of all available trials that matched these criteria. No duplicate registrations were detected at the time.

**Table 2:**
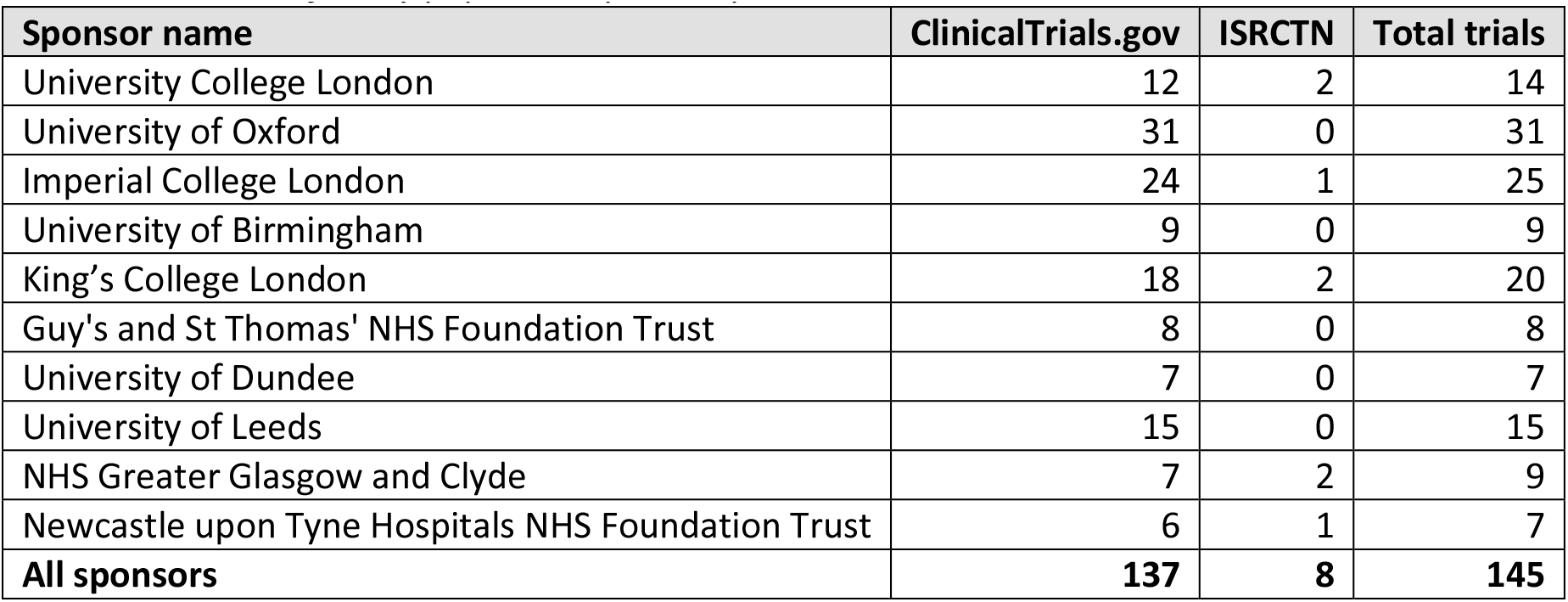
Overview of study population prior to publication searches

| Sponsor name | ClinicalTrials.gov | ISRCTN | Total trials |
| --- | --- | --- | --- |
| University College London | 12 | 2 | 14 |
| University of Oxford | 31 | 0 | 31 |
| Imperial College London | 24 | 1 | 25 |
| University of Birmingham | 9 | 0 | 9 |
| King's College London | 18 | 2 | 20 |
| Guy's and St Thomas' NHS Foundation Trust | 8 | 0 | 8 |
| University of Dundee | 7 | 0 | 7 |
| University of Leeds | 15 | 0 | 15 |
| NHS Greater Glasgow and Clyde | 7 | 2 | 9 |
| Newcastle upon Tyne Hospitals NHS Foundation Trust | 6 | 1 | 7 |
| **All sponsors** | **137** | **8** | **145** |

The final cohort consists of 145 interventional clinical trials run by ten major non-commercial UK sponsors that are registered on ClinicalTrials.gov or ISRCTN and that were completed or terminated during 2017. These trials had a combined (actual or planned) enrolment of 34,102 patients.

### Publication search strategy

The lead author recruited volunteers to perform the publication searches and provided them with a literature search guide detailing a 3-step process for locating publications in scientific journals, the literature and/or trial registries. Involving volunteers helped to raise awareness of publication bias and research waste within the clinical community, and gave medical students and early career researchers experience in conducting literature searches and participating in meta-research projects. The search strategy is a simplified version of strategies commonly used in comparable academic meta-research studies. It is described in detail in the study protocol.

During the initial data extraction, 25/145 trials were identified as having tabular summary results available on ClinicalTrials.gov; these were marked as “reported” prior to the literature search.

In November 2022, volunteers searched for publications for the remaining 120/145 trials, entering links to trial results into an online spreadsheet. As per protocol, tabular summary results posted onto clinical trial registries, articles published in peer-reviewed journals and PhD theses were classified as publications. Conference abstracts, posters, presentation slides, and other documents containing trial outcomes were classified as ‘grey literature’. The lead researcher reviewed all publications to verify that they contained trial outcomes and had been accurately classified. As the aim was to capture all publications, he also performed some supplementary searches.

### Data validation with sponsors

In December 2022, the lead researcher contacted the press offices of all sponsors by email with a list of their trials for which no results had been located. Sponsors were invited to (a) flag any relevant publications that the study team may have overlooked (based on a dataset shared with sponsors), and to (b) provide a short on-the-record statement on their clinical trial reporting policies and plans.

The emails identified the study as being run by TranspariMED. 9/10 sponsors (90%) responded. In January-February 2023, the lead researcher obtained the remaining data from the only non-responsive sponsor (University of Oxford) through a Freedom of Information request.

The 100% sponsor response rate made the originally planned second round of literature searches superfluous (protocol deviation).

On 01 March 2023, the lead researcher performed a final registry and literature search using trial ID numbers only to capture possible recent publications; no additional trial results were found.

The study cohort size remained unchanged at 145 trials as no trial had been identified as ‘ongoing’ or ‘withdrawn’. Some trials were identified as having been registered on more than one registry; their alternative registry numbers were added to the spreadsheet.

### Protocol registration, ethics approval, funding and data availibility

After compilation of the trial cohort, the study protocol was registered on OSF (https://osf.io/rh3m9) prior to the start of publication searches. A UK Health Research Authority NHS REC ethics waiver was secured on 01 November 2022.

This study was funded by HealthSense UK (formerly HealthWatch UK), a UK registered charity, in 2019. Study startup was delayed by several years due to the pandemic. The research protocol departs from the original research proposal submitted to HealthSense UK in order to maximise the study’s relevance to current policy making.

The outcomes of this study are reported in line with the STROBE guideline for cohort studies.

The study protocol, dataset, literature search guide, ethics waiver and sponsors’ responses are publicly available on GitHub (https://github.com/TillBruckner/UKtrials).

## RESULTS

The study hypothesis that the results of some clinical trials in the cohort were never made public was confirmed.

In total, 116/145 trials (80%) had reported results on a registry, in the academic literature, or in a PhD thesis. The outcomes of 11/145 trials (8%) had been reported in the grey literature. Results for 18/145 trials (12%) completed during 2017 remained completely unpublished as of early March 2023.

**Table 3:** Number and percentage of trials that have not fully reported results

|  | Number | Percentage |
| --- | --- | --- |
| Results fully reported | 116 | 80% |
| Results not fully reported* | 29 | 20% |
| Total trials | 145 | 100% |

*\* Note: Includes grey literature publications*

In total, 637 people were enrolled in the 18 clinical trials that remained completely unreported. Assuming an average cost per trial of £500,000, the total aggregate cost of those 18 trials was £9 million.

Sponsors indicated that they still planned to make public the results of 14/18 unreported trials. For the remaining 4/18 trials, sponsors indicated that there was no data of value to publish due to early termination of the trial (3/18 trials) or data quality issues (1/18 trials). A total of 133 people participated in these ‘written off’ trials.

**Table 4:** Unreported clinical trials and future publication plans

| Sponsor | Trial ID | People | Sponsor plans to publish |  |
| --- | --- | --- | --- | --- |
| Guy's and St Thomas<sup>1</sup> | NCT02763631 | 21 | YES | Sponsor plans to audit entire portfolio in near future |
| Guy's and St Thomas | NCT02765360 | 22 | YES |  |
| Guy's and St Thomas | NCT02863835 | 50 | YES |  |
| Guy's and St Thomas | NCT02952625 | 8 | YES |  |
| Guy's and St Thomas | NCT03258060 | 1 | YES |  |
| Guy's and St Thomas | NCT03609970 | 36 | YES |  |
| Imperial College London | NCT02874820 | 2 | **NO** | **No data of value** |
| King's College London | ISRCTN71271888 | 20 | YES | "in due course" |
| NHS Greater Glasgow & Clyde | ISRCTN14022536 | 60 | YES | Have contacted PI |
| University College London | NCT02261753<sup>2</sup> | 3 | **NO** | **No data of value** |
| University of Birmingham | NCT02426515 | 63 | YES | Have contacted PI |
| University of Birmingham | NCT02443896 | 100 | YES | Have contacted PI |
| University of Birmingham | NCT02605291 | 13 | YES | PI has left, will follow up |
| University of Dundee | NCT02984293 | 18 | YES | Submitted to journal |
| University of Leeds | NCT02725775 | 90 | YES | PI intends to publish |
| University of Leeds | NCT04154852<sup>3</sup> | 22 | **NO** | **No data of value** |
| University of Oxford | NCT01640587 | 76 | **NO** | **"study abandoned"** |
| University of Oxford | NCT02495246 | 32 | YES | Planned for 2023 |

## DISCUSSION

### Summary of findings

This study found a full publication rate of 80% for this cohort of clinical trials after a follow-up period of 5-6 years. Including grey literature, the publication rate is 88%. Publication rates increased slightly as a result of our outreach to sponsors. They are far higher than what comparable studies have found for trial cohorts in other countries. Overall, sponsors displayed a very high willingness to pursue the publication of currently unreported clinical trial results.

### Strengths and weaknesses

The findings of this study cannot be generalised to all UK clinical trials for multiple reasons. The study cohort was small, CTIMPs were underrepresented, large sponsors tend to have higher than average reporting rates, and at least one sponsor appears to have reported several results following our outreach. The actual cost of research waste may be significantly higher or lower than our rough estimate of £9 million. As with similar studies, our methodology was unable to capture unregistered trials. A key strength of this study is that we succeeded in verifying the publication status of every clinical trial with its responsible sponsor.

### Responses by sponsors

Sponsors’ reaction to our outreach was extremely positive overall. Of note, Imperial College London uploaded several trial results onto ClinicalTrials.gov shortly after being contacted. Guy’s and St Thomas’ NHS Foundation Trust were already planning to conduct a full audit of trials conducted over the past five years. All sponsors with results still outstanding initiated steps to encourage future publication. Note that there is currently no legal or regulatory requirement for UK sponsors to do any of the above for legacy non-CTIMP trials. Sponsor responses are archived on GitHub.

### Study impact on reducing research waste

Our study was not geared towards measuring its potential impact on reducing research waste. Our methodology neither included a control group nor longer-term follow-up. However, at a minimum, our study accelerated the reporting of outcomes of several trials sponsored by Imperial College London.

Several sponsors’ responses indicated that they had not been systematically tracking publication status within their non-CTIMP legacy portfolios and therefore had not been aware of their legacy unreported trials. Sponsors’ responses suggest that it is likely that our outreach will in future lead to the publication some clinical trial results that would otherwise have ended up as research waste. Our dataset is public and we encourage other researchers to follow up this cohort in future.

### Policy implications

The sponsors in our cohort only started systematically uploading CTIMP results onto the EUCTR registry in 2018-2019. Today, all ten sponsors have an excellent CTIMP reporting record, illustrating that locating, analysing and disclosing legacy outcome data, including data more than a decade old, is generally possible.

Replicating this project on a larger scale would appear to be a highly cost-effective way to expand the global store of medical knowledge. The entire budget of this project was a small fraction of the cost of a typical trial. TranspariMED is currently working on scaling up the approach to identify and follow up on all unreported trials involving UK patients listed on ISRCTN.

Our findings suggest that the UK Health Research Authority could improve future clinical trial reporting by fine-tuning its policies in three areas.

First, 11 trials (8% of the cohort) had published outcomes only in what this study defined as ‘grey literature’. Not all disclosure formats are equal. For example, Tweets and press releases are widely considered to not constitute adequate disclosure. Going forward, the HRA should clarify what does, and does not, constitute an acceptable disclosure format. The HRA should require all results to be made public on trial registries, as recommended by the World Health Organisation, as registry reporting has significant advantages over other publication formats.^9^

Second, some results had been made public but were not findable in practice. For example, the results of one trial (NCT02794389) had been published in a PhD thesis that did not contain the trial ID number. The HRA should encourage sponsors to include the trial ID number(s) in any document containing trial outcomes. The HRA should require sponsors to add a hyperlink to every outcome publication to the relevant registry entry.

Third, sponsors had no plans to publish the results of 4 trials (3% of the cohort) because they perceived the data to have no value. Two of these trials recruited only 2-3 participants each, and a third (NCT04154852 / EUCTR2008-004877) had obtained an MHRA reporting waiver due to data quality issues. In these three cases, allocating finite resources to making results public is unlikely to be in the public interest. However, in the case of the fourth trial (NCT01640587), the sponsor stated that “[t]his study was abandoned because of recruitment problems,” but at that point, 76 people had already been enrolled. The HRA should establish a pathway allowing sponsors to apply for HRA permission to ‘write off’ a trial’s results in exceptional circumstances, provided that such waivers are noted in the relevant registry.

## Data Availability

All data produced are available online at Github.

https://github.com/TillBruckner/UKtrials

